# Impact of remdesivir on 28 day mortality in hospitalized patients with COVID-19: February 2021 Meta-analysis

**DOI:** 10.1101/2021.03.04.21252903

**Authors:** Robert Robinson, Vidhya Prakash, Raad Al Tamimi, Nour Albast, Basma Al-Bast, Elizabeth Wieland, Carlos Garcia

## Abstract

**Background:** The COVID-19 pandemic has stimulated worldwide investigation into a myriad of potential therapeutic agents, including antivirals such as remdesivir. The first RCT reporting results on the impact of remdesivir on COVID-19 in a peer reviewed journal was the ACTT-1 trial published in November, 2020. The ACTT-1 trial showed more rapid clinical improvement and a reduced risk of 28-day mortality in patients who received remdesivir.

This study is a meta-analysis of peer reviewed RCTs aims to estimate the association of remdesivir therapy compared to the usual care or placebo on all-cause mortality in hospitalized patients with COVID-19. Software based tools to accelerate the analysis process.

**Methods:** Meta-analysis of peer reviewed RCTs comparing remdesivir to usual care or placebo. The protocol for this meta-analysis was registered and published in the PROSPERO database (CRD42021229985) on February 5, 2021.

**Results:** Four English language RCTs were identified, including data from 7,333 hospitalized patients worldwide using remdesivir in COVID-19 positive patients.

Meta-analysis of all identified RCTs showed no difference in survival in patients who received remdesivir therapy compared to usual care or placebo. The random effects meta-analysis has a summary odd ratio is 0.89 (95% CI 0.65-1.21, p = 0.30). Considerable variability in the severity of illness is noted with the rates of IMV at the time of randomization ranging from 0% to 27%.

**Conclusions:** This meta-analysis of randomized controlled trials published in peer-reviewed literature by February 1, 2021 did not show reduced mortality in hospitalized patients with COVID-19 who received remdesivir. Further research is needed to clarify the role of remdesivir therapy in the management of COVID-19.

## Introduction

Coronavirus disease 2019 (COVID-19) first reported in Wuhan, China in December 2019 with symptoms of severe viral pneumonia. The novel virus also known as severe acute respiratory syndrome coronavirus 2 (SARS-CoV-2) has since become a pandemic spreading through an immunologically naïve population.^1^ To date, over 109 million people have been infected and 2.4 million people have died worldwide.^2^ Previous outbreaks of coronaviruses (CoV) include severe acute respiratory syndrome (SARS) in 2002 and Middle East respiratory syndrome (MERS) in 2012.^3^ Patient care for human coronaviruses (HCoV) infections have focused on symptom relief and supportive care.^3^

Coronaviruses are difficult to treat because the virus has a large breadth of antigenic diversity. ^1^ CoV are positive, single-stranded, RNA viruses that are inherently error prone during viral replication leading to an increase in diversity among the virus family. ^1,4^ This diversity explains the wide spectrum of disease endpoints, ranging from self-limiting infections to severe infections that may lead to death.^3,4^

The COVID-19 pandemic has stimulated worldwide investigation into a myriad of potential therapeutic agents. Existing knowledge about antiviral medication in combating SARS and MERS epidemics served as a starting point for many potential treatments for COVID-19.^3^ Some of the antiviral medications being studied in randomized controlled trials were developed for other therapeutic reasons, but showed benefit in combating CoV, including remdesivir.^5^ Remdesivir was developed as a potential treatment for Ebola virus and has been shown to inhibit SARS and MERS replication in vitro and in mouse models.^1^ In addition to antiviral activity against SARS, MERS, and Ebola, Remdesivir has been shown to inhibit a broad spectrum on RNA viruses including RSV, Junin virus, and other coronaviruses that lead to upper respiratory disease.^3,5^

Remdesivir is an adenosine nucleotide analog that works as an RNA dependent-RNA polymerase (RDRP) inhibitor and blocks viral RNA synthesis.^3^ Due to the amino acid sequence similarity between the SARS-CoV and SAR-CoV-2’s RDRP, there was basis to assume that remdesivir could inhibit SARS-CoV-2 replication similar to its inhibition of SARS-CoV in previous studies.^1,3^ The RDRP is an essential component of the coronavirus lifecycle and is highly conserved; therefore, drugs targeting this mechanism are promising for COVID-19 mitigation.^6^

Observational and randomized controlled trials (RCTs) were quickly initiated to evaluate the impact of antivirals such as remdesivir on COVID-19. Based on in-vitro studies demonstrating remdesivir activity against SARS-CoV-2, the first positive COVID patient in the U.S was administered remdesivir for compassionate use when his clinical status worsened. Within days after administration the patient’s need for oxygen supplementation decreased and he began showing signs of clinical improvement.^7^

Shortly after, Gilead Sciences began an observational study to further investigate remdesivir for compassionate use in patients with SARS-CoV-2 infections. The study analyzed 53 patients that had received remdesivir from January 25^th^ through March 7th.

The results showed an overall mortality rate of 13% (7/53) after completion of remdesivir treatment. Patients that required invasive ventilation had a higher mortality rate of 18%. However, 68% (36/53) of patients with severe COVID-19 that received remdesivir clinically improved in this study.^8^ By comparison, an early study in Wuhan, China in patients that did not receive remdesivir showed an overall mortality rate of 22% and a mortality rate of 66% for patients who were requiring either invasive ventilation or ICU admission.^9^ While these observations suggested that remdesivir could be clinically beneficial, further studies were needed in order to assess its efficacy as well as patient safety.

The Adaptive COVID-19 Treatment Trial-1 (ACTT-1) was an early study of remdesivir for treatment of SARS-CoV-2. ACTT-1 showed promising preliminary results in recovery time for hospitalized and reduced need for intubation.^10^ Due to this, remdesivir became the first drug to receive emergency use authorization from the FDA for the treatment of hospitalized patients with COVID-19.^10^ However, once the control and experimental groups were stratified by initial respiratory support, there was evidence that the remdesivir group had by chance received less severe patients compared to the placebo group.^11^ Analysis and correlation of other available RCT’s is needed in order to assess remdesivir’s effect on length of hospital stay, reduction in need for intubation, and mortality benefit.

This meta-analysis of RCTs published in peer reviewed literature is designed to estimate the association of remdesivir therapy compared to the usual care or placebo on 28-day mortality in hospitalized patients with COVID-19. This meta-analysis utilized software-based tools to accelerate the analysis process.

## Methods

Data management and statistical analysis was conducted with R version 4.0.2. Meta-analysis calculations and plots were generated using the *meta* (version 4.15-1) and *metafor* (version 2.4-0) packages for R.

The protocol for this meta-analysis was registered and published in the PROSPERO database (CRD42021229985) on February 5, 2021. A PRISMA checklist is included in this document.

### Identification of trials

Trials were identified by searching MEDLINE using the search terms *COVID-19* and *remdesivir*. The results of these searches were exported and filtered via the RobotSearch tool by Vortext Systems to identify randomized controlled trials.^12^ The filtered results were reviewed for inclusion criteria and evaluated for bias via the RobotReviewer tool that applies the Cochrane Risk of Bias (RoB) tool to the full text of articles.^13^ The bibliographies of included trials were also searched for references to additional trials.

Inclusion criteria for this study were: RCTs comparing remdesivir to other treatments or placebo in hospitalized adults with COVID-19 infections published by January 1, 2021 in an English language MEDLINE-indexed peer reviewed journal.

### Meta-analysis

A random-effects meta-analyses using the Paule-Mandel estimate of heterogeneity was used to determine odds ratios. The Hartung-Knapp adjustment was used to address uncertainty in the estimates of between-study variance in the random-effects meta-analysis.

Inconsistency in associations were evaluated using the I^2^ statistic and the Cochran Q statistic. Exact p values are reported.

Odds ratios and 95% confidence intervals were plotted for each trial to evaluate the association between remdesivir therapy compared with usual care or placebo to the outcome measure.

Meta-analysis of the trials reporting data on critically ill patients included all trials where intensive care unit (ICU) admission was an eligibility criteria and the data on the patients who were on invasive mechanical ventilation (IMV) at randomization in all other trials.

Meta-analysis of trials reporting data on non-critically ill patients excluded all trials where ICU admission was an eligibility criteria and the data on the patients who were not on IMV at randomization in all other trials.

### Outcomes

The primary outcome evaluated for this meta-analysis was all cause mortality up to 30 days after randomization. Shorter timeframes were acceptable if 30-day survival data was not available.

## Results

The search strategy identified 4 English language RCTs that had results published in peer reviewed literature (Figure 1). They included data of 7,333 hospitalized patients in numerous centers from all over the world. 3826 patients were randomized to receive remdesivir compared with 3507 patients were randomized to the control groups. Remdesivir was either compared to placebo (3 RCTs) or the usual care (1 RCT).

**Figure 1.**
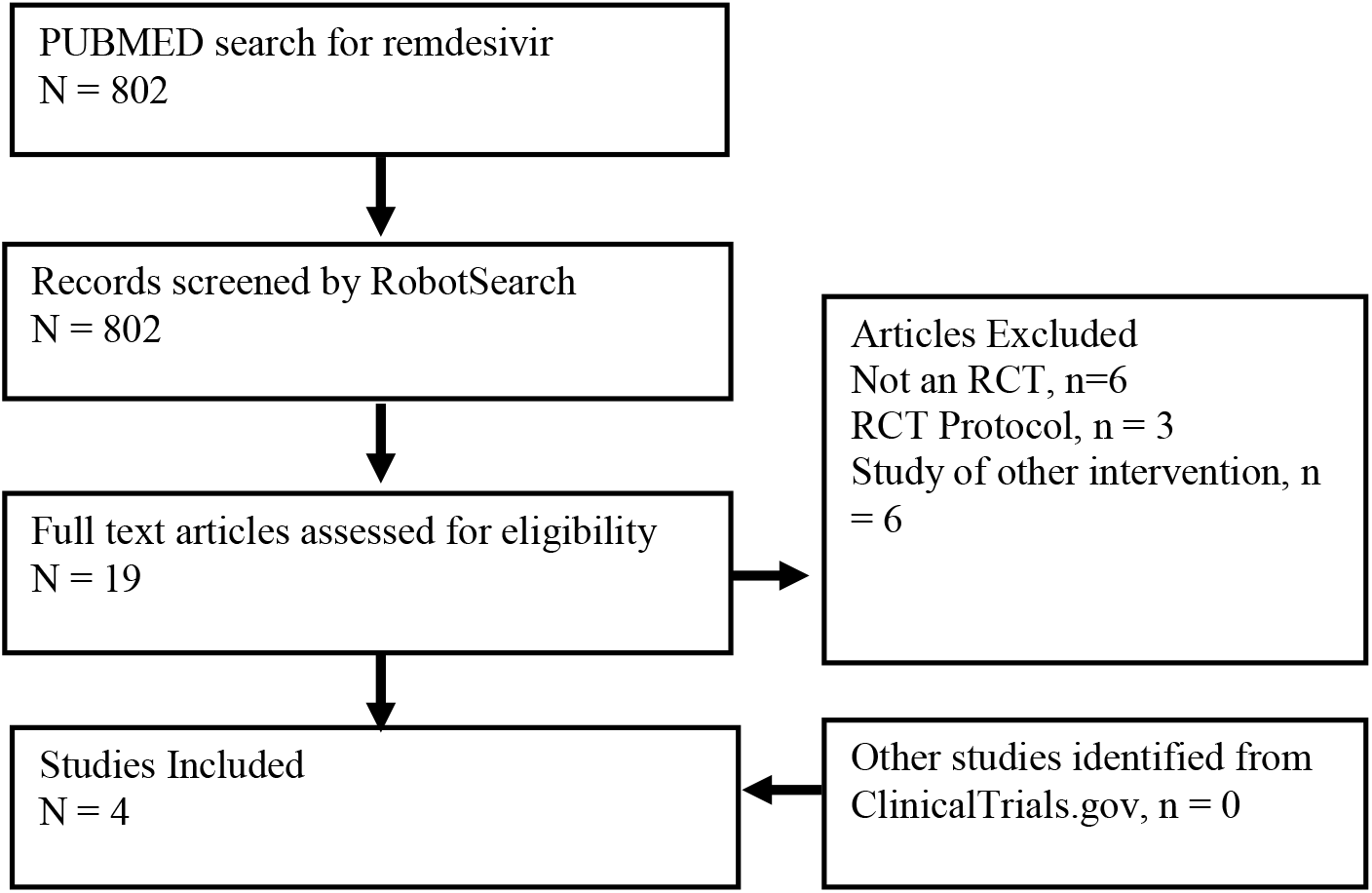
Article Selection Flowchart.

### ACTT-1 Trial

The ACTT-1 trial (NCT04280705) enrolled and randomized 1062 patients to receive either remdesivir or placebo at 60 trial sites worldwide.^10^ The median age in this study was 59 years, 36% of participants were women and 27% were on IMV at the time of randomization. The risk of bias in this placebo controlled RCT was assessed to be low. This study showed no difference in 28-day mortality between the intervention and control groups (11% vs. 15%, odds ratio = 0.73; 95% CI = 0.52 to 1.03, p = non-significant, Figure 2).

**Figure 2.**
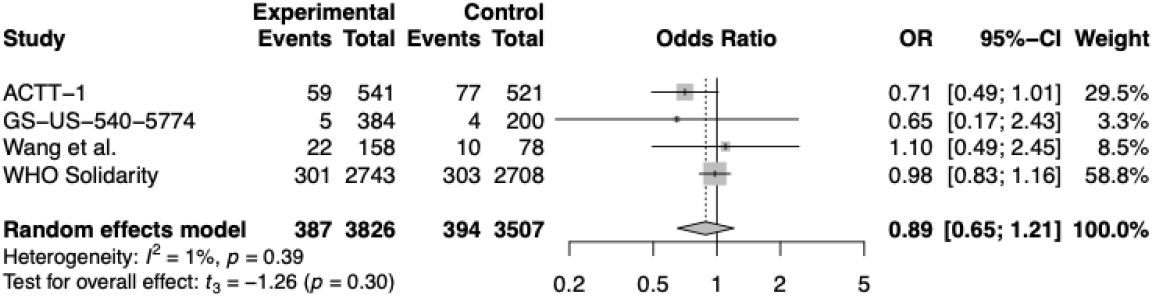
Association Between remdesivir and All-Cause Mortality in All Identified RCTs.

### GS-US-540-5774

The GS-US-540-5774 (NCT04280705) enrolled and randomized 584 patients to receive either remdesivir or placebo at 105 trial sites worldwide.^14^ The median age in this study was 57 years, 39% of participants were women and no participants were on IMV at the time of randomization. The risk of bias in this placebo controlled RCT was assessed to be moderate because treatment assignment was not blinded. This study showed no difference in 11-day mortality between the intervention and control groups (1% vs. 2%, odds ratio = 0.65, 95% CI = 0.17 to 2.43, p = non-significant, Figure 2).

### Wang et al. Trial

The Wang et al. trial (NCT04257656) enrolled and randomized 237 patients to receive either remdesivir or placebo at 10 hospitals in China.^15^ The median age in this study was 66 years, 41% of participants were women, and less than 1% were on IMV at the time of randomization. The risk of bias in this placebo controlled RCT was assessed to be low. This study showed no difference in 28-day mortality between the intervention and control groups (14% vs. 13%, odds ratio = 1.10, 95% CI = 0.49 to 2.45 p = non-significant, Figure 2).

### WHO Solidarity Trial

The WHO Solidarity trial (NCT04315948) enrolled and randomized 2750 patients to receive either remdesivir or usual care at 405 hospitals in 30 countries.^11^ The most common age range in this study was 50-69 years of age (47%), 37% of participants were women and 9% were on IMV at the time of randomization. The risk of bias in this RCT was assessed to be moderate because treatment assignment was not blinded. The full protocol for this study was published separately.^16^ This study showed no difference in 28-day mortality between the intervention and control groups (11% vs. 11%, odds ratio = 0.98, 95% CI = 0.83 to 1.16, p >0.10, Figure 2).

### Association between remdesivir and mortality in all identified RCTs

There were 387 deaths in the 3826 patients randomized to receive remdesivir compared with 394 deaths in the 3507 patients randomized to the control groups. Critically ill patients and patients who were not critically ill were included in this meta-analysis. The random effects meta-analysis has a summary odd ratio is 0.89 (95% CI 0.65-1.21, p = 0.30, Figure 2). Heterogeneity assessment shows an I^2^ of 1% (p = 0.39). Considerable variability in the severity of illness is noted with the rates of IMV at the time of randomization ranging from 0% to 27%.

The GS-US-540-5774 trial which randomized 584 participants reports mortality at 11 days, which is a much shorter time frame than the 28 days for the other 3 trials. Excluding this trial for a random effects meta-analysis shows a summary odds ratio of 0.89 (95% CI 0.55-1.46, p = 0.43, Figure 3), which is very similar to the meta-analysis including data from all identified trials.

**Figure 3.**
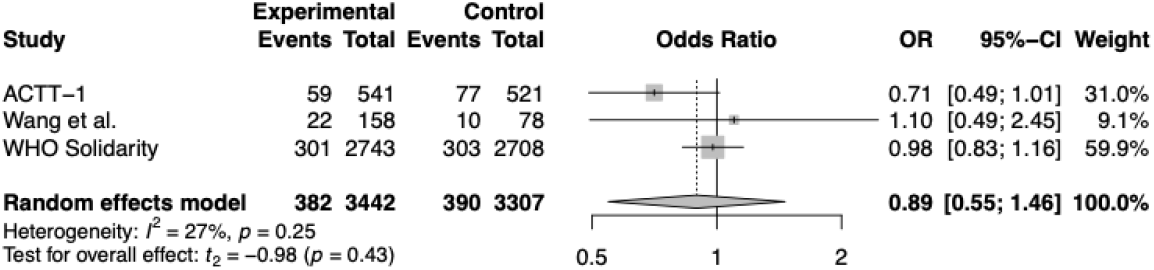
Association Between remdesivir and 28-day All-Cause Mortality in RCTs.

## Discussion

This meta-analysis of RCTs published in peer-reviewed literature by January 1, 2021 showed no statistically significant difference in all-cause mortality in patients with COVID-19 who received remdesivir compared with placebo or the usual care.

A meta-analysis of RCTs published by October 25, 2020 identified 3 peer reviewed trials and no impact on mortality (odds ratio 0.77, 95%CI 0.56-1.06, p = 0.108).^17^ All the trials identified in the earlier meta-analysis were found and included in this meta-analysis.

The 28-day mortality was reported in all identified trials except for the GS-US-540-5774 trial, which reported 11-day mortality only. This study analyzed the results including and excluding the GS-US-540-5774 trial, and it showed similar results in both circumstances.

Notably, mortality was not the primary outcome in all of the identified trials except for the WHO Solidarity trial. Instead, time to recovery and clinical improvement were studied as primary outcomes. All of the identified trials reveled no statistically significant difference in all-cause mortality. However, the Adaptive COVID-19 Treatment Trial (ACTT-1), which is the earliest RCT to study remdesivir for the treatment of COVID-19, showed a shorter time for recovery and better clinical status. This trial changed its primary outcome to evaluate the participants by day 29 instead of day 15 due to new data suggesting that COVID-19 has a longer clinical course than previously anticipated.

The findings of this meta-analysis along with taking into consideration the potential adverse effects of remdesivir therapy, question the justification of its use in the management of hospitalized patients with COVID-19.

This study has several important limitations. The focus on English language publication in peer reviewed literature may under-represent the full spectrum of research in this rapidly evolving field. Remarkably, these 4 trials were conducted and published within 9 months of the World Health Organization declaration of the COVID-19 pandemic on March 11, 2020.

The literature search and assessment methods used for this study is computer assisted, which differs from traditional meta-analysis techniques that are labor and time consuming. This strategy has the potential to introduce bias in the identification of RCTs. However, this methodology did produce similar results to other systematic reviews and meta-analyses on the same topic.^17^

## Conclusions

This meta-analysis of randomized controlled trials published in peer reviewed literature by January 1, 2021 reveals no significant impact of remdesivir therapy on all-cause mortality in hospitalized patients with COVID-19. There is evidence that may suggest some benefit of remdesivir in the clinical course of the disease and recovery. However, further research is needed to clarify its role in the management of COVID-19.

## Data Availability

Data included in manuscript and in original publications

---

**Table 1.** Included RCTs

| Table 1. Included RCTs |  |  |  |  |
| --- | --- | --- | --- | --- |
|  | ACTT-1 | GS-US-540-5774 | Wang et al. | WHO Solidarity |
| ClinicalTrials.gov ID | NCT04280705 | NCT04280705 | NCT04257656 | NCT04315948 |
| Patients |  |  |  |  |
| Number | 1062 | 584 | 237 | 2750 |
| Age (Median) | 59 years | 57 years | 66 years | 50-69 years (median) |
| Women | 36% | 39% | 41% | 37% |
| Confirmed COVID-19 | 100% | 100% | 100% | 100% |
| IMV at enrollment | 27% | 0% | <1% | 9% |
| Eligibility criteria |  |  |  |  |
| Known/Suspected COVID-19 | Yes | Yes | Yes | Yes |
| Acute respiratory failure |  | Yes | Yes |  |
| Pulmonary infiltrates on imaging study |  | Yes | Yes |  |
| Intervention - Remdesivir |  |  |  |  |
| Dose | Remdesivir 200mg IV Day 1 then 100mg IV Daily for 9 additional days | Remdesivir 200mg IV Day 1 then 100mg IV Daily for 4 or 9 additional days | Remdesivir 200mg IV Day 1 then 100mg IV Daily for 9 additional days | Remdesivir 200mg IV Day 1 then 100mg IV Daily for 9 additional days |
| Control |  |  |  |  |
| Outcome | 28 day mortality | 11 day mortality | 28 day mortality | 28 day mortality |
| Setting | Hospital | Hospital | Hospital | Hospital |
| Location | UK, EU, Korea, Mexico, Spain, Japan | US, EU, Asia | China | Worldwide |
| Risk of bias |  |  |  |  |
| Random sequence generation | Yes | Yes | Yes | Yes |
| Allocation concealment | Yes | No | Yes | No |
| Blinding of participants and personnel | Yes | Yes | Yes | Yes |
| Blinding of outcome assessment | Yes | Yes | Yes | Yes |

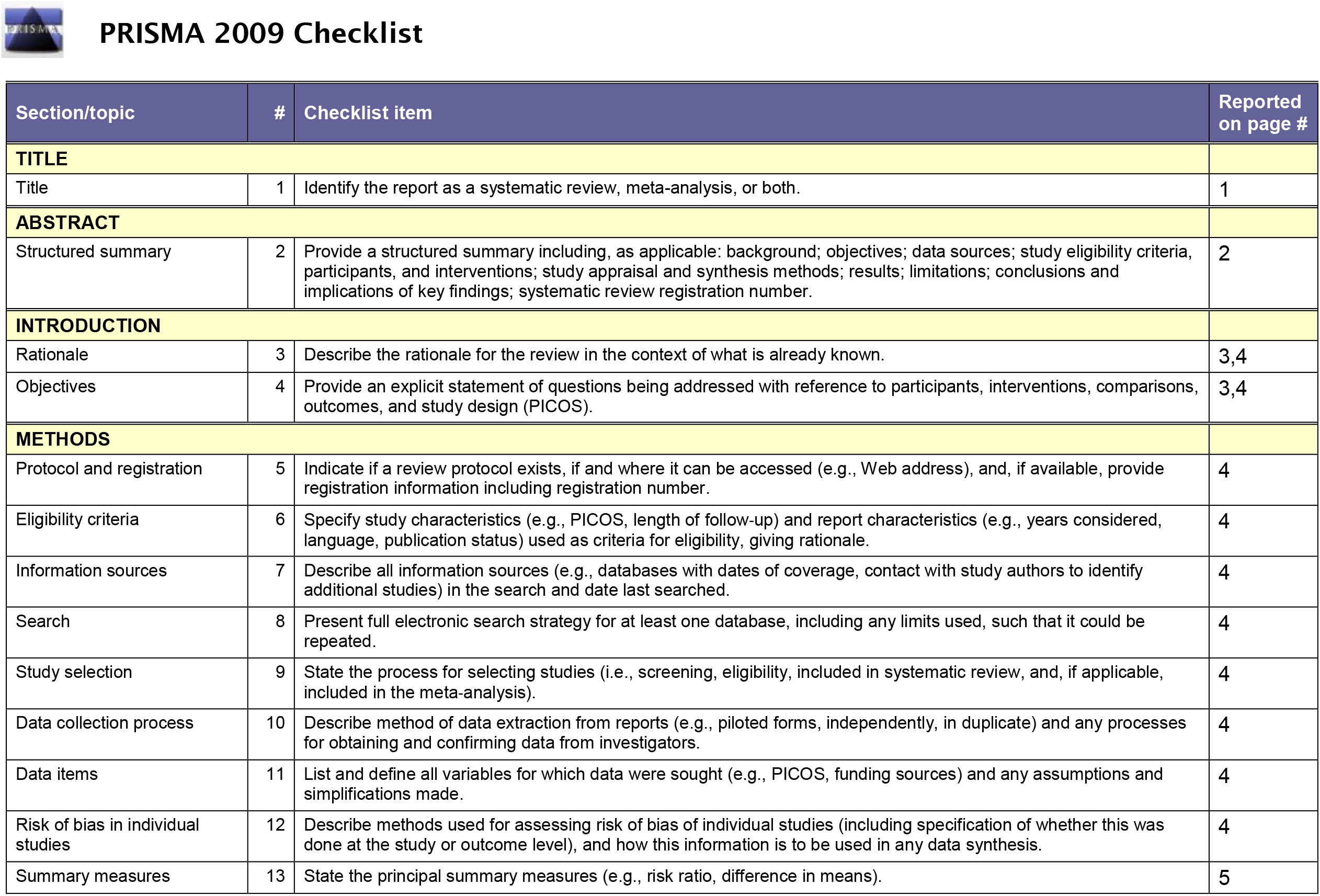
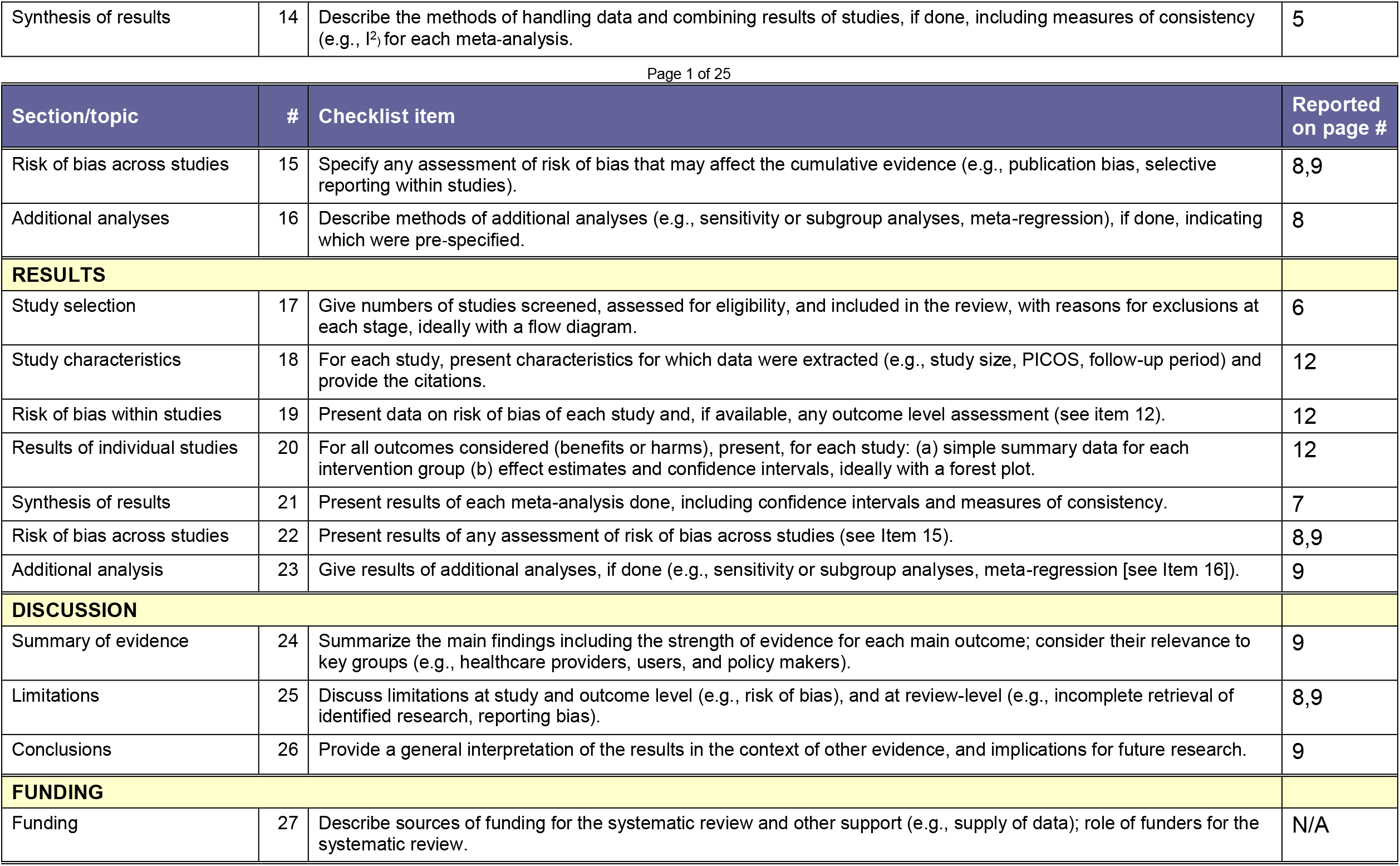
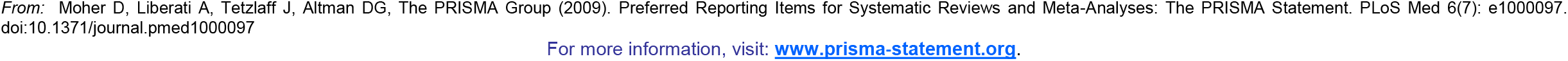

